# An immune correlate of SARS-CoV-2 infection and severity of reinfections

**DOI:** 10.1101/2021.11.23.21266767

**Authors:** Hannah E. Maier, Angel Balmaseda, Sergio Ojeda, Cristiam Cerpas, Nery Sanchez, Miguel Plazaola, Harm van Bakel, John Kubale, Roger Lopez, Saira Saborio, Carlos Barilla, PSP Study Group, Eva Harris, Guillermina Kuan, Aubree Gordon

## Abstract

**Background:** An immune correlate of protection from SARS-CoV-2 infection is urgently needed.

**Methods:** We used an ongoing household cohort with an embedded transmission study that closely monitors participants regardless of symptom status. Real-time reverse-transcription polymerase chain reaction (RT-PCR) and Enzyme-linked immunosorbent assays (ELISAs) were used to measure infections and seropositivity. Sequencing was performed to determine circulating strains of SARS-CoV-2. We investigated the protection associated with seropositivity resulting from prior infection, the anti-spike antibody titers needed for protection, and we compared the severity of first and second infections.

**Results:** In March 2021, 62.3% of the cohort was seropositive. After March 2021, gamma and delta variants predominated. Seropositivity was associated with 69.2% protection from any infection (95% CI: 60.7%-75.9%), with higher protection against moderate or severe infection (79.4%, 95% CI: 64.9%-87.9%). Anti-spike titers of 327 and 2,551 were associated with 50% and 80% protection from any infection; titers of 284 and 656 were sufficient for protection against moderate or severe disease. Second infections were less severe than first infections (Relative Risk (RR) of moderated or severe disease: 0.6, 95% CI: 0.38-0.98; RR of subclinical disease:1.9, 95% CI: 1.33-2.73).

**Conclusions:** Prior infection-induced immunity is protective against infection when predominantly gamma and delta SARS-CoV-2 circulated. The protective antibody titers presented may be useful for vaccine policy and control measures. While second infections were somewhat less severe, they were not as mild as ideal. A strategy involving vaccination will be needed to ease the burden of the SARS-CoV-2 pandemic.

---

The course of the current SARS-CoV-2 pandemic will be determined by the quality and durability of protective immunity induced by prior infection or vaccination, as well as the severity of illness in individuals with some level of immunity.^1-3^

As the SARS-CoV-2 pandemic continues in its second year, vaccine availability is still sorely limited, particularly in low- and middle-income countries, and an immune correlate of protection is urgently needed.^4^ Such an immune correlate of protection would inform vaccine policy, allow more rapid development of vaccines, and could guide targeting of at-risk populations for vaccination—including when and how often to boost.

Recent vaccine efficacy studies have estimated the levels of vaccine-induced antibodies needed for protection.^5,6^ However, with the increased circulation of SARS-CoV-2 variants, studies are needed to evaluate protection against these variants. Immune correlates are also needed for natural infection-induced protection and for children, and they need to be stratified by severity of infection.^7^

As of November 8, 2021, ∼40% of the global population was fully vaccinated, with 66% of the population in high-income countries, but only 2% of that in low-income countries fully vaccinated.^8^ Yet, much of the world’s population has been infected,^8^ making it critical to understand the degree of protection achieved in previously infected persons and populations, particularly in low- and middle-income countries. An immune correlate of natural-immunity induced protection and a better understanding of the severity of second infections will help inform models estimating the future burden and will continue to guide mitigation strategies.^2^

We used data from an existing household cohort study of 2,353 individuals 0 to 94 years of age.^9^ We previously published on the first wave of SARS-CoV-2 in this cohort, reporting that ∼56% of the cohort was seropositive in October 2020.^9^ Here we focus on infections that occurred during the second pandemic wave, after March 2021, in which gamma and delta variants predominated. We measure the protection from SARS-CoV-2 infection during the second wave associated with immunity induced by prior infection, as well as the “ absolute correlate” or protective threshold^4^ of antibodies needed for 50% and 80% protection. Finally, we compare the severity of first and second infections during the second wave.

## METHODS

### The Household Influenza Cohort Study (HICS)

The Nicaraguan HICS is an ongoing prospective study of influenza in households that are free of disease at baseline. Located in district II of Managua, Nicaragua, the HICS started in 2017 and was expanded in February 2020 to include the study of SARS-CoV-2 infection and disease. At the first indication of any illness, participants are requested to report to the study health center, where they are provided with their primary care. Blood samples for serology are collected annually in March-April, and an extra midyear sample was collected for those who consented to it in October 2020. A transmission sub-study is nested within HICS, in which participants are monitored closely and tested regardless of symptoms once a SARS-CoV-2 case is detected in their household (see supplement for more details). This study was approved by the institutional review boards at the Nicaraguan Ministry of Health and the University of Michigan (HUM00119145 and HUM00178355). Informed consent or parental permission was obtained for all participants. Assent was obtained from children aged ≥6 years.

### Laboratory Assays

Real-time reverse-transcription polymerase chain reaction (RT-PCR) was performed according to the protocol from Chu et al.^10^ Enzyme-linked immunosorbent assays (ELISAs) were run on paired serum samples (current vs baseline) with a protocol adapted from the Krammer laboratory.^11^ The SARS-CoV-2 spike receptor binding domain (RBD) and spike proteins for ELISAs were produced in single batches at the Life Sciences Institute at the University of Michigan. For ELISAs, spike RBD was used for screening (positive/negative) and spike was used for titers. The limits of detection for endpoint titers were 100 (lower) and 6,400 (upper). All RT-PCR and most ELISAs were performed at the Nicaraguan National Virology Laboratory, with a minority of 2020 and 2019 annual samples processed at the University of Michigan. Sequencing information is presented in the supplement.

### Seropositivity, RT-PCR-positive episodes, and severity

ELISA seropositivity in October 2020 and March 2021 was defined as shown in fig. S1 Seroconversion was measured in most cases, by comparing the current to the baseline (March 2020 and March 2019) ELISA results, but we use the term seropositivity due to a small subset of participants not having baseline samples. Potentially cross-reactive antibodies—those that were positive at baseline with less than a 4-fold rise in titer—and indeterminate ELISA results were counted as missing.

RT-PCR-positive episodes were considered separate episodes if they were ≥60 days apart. To assess the difference in severity between first and second SARS-CoV-2 infections, the data were not limited to the subset of participants with ELISA results. Second infections were all RT-PCR-positive after March 1, 2021, but first infections could have been a first RT-PCR-positive infection or prior seropositive status (seropositive in October 2020 or March 2021, figs. S1 and S2).

Symptom data from multiple sources was used to classify infection severity (see supplement for more details).

### Analysis

All analyses were performed in R version 4.1.1.^12^ using the tidyverse^13^. Participant age was calculated on March 1, 2021, near the 2021 annual sample collection. A Poisson distribution was used to calculate 95% confidence intervals (CIs) for attack rates, risk ratios, and percent protection (1 – risk ratio). For those without numeric titers, the results were classified as follows: if screened RBD negative, titer was set to 5 (n=817); if RBD positive but no titer available (n=1), titer was set to 20; if titer was <100 (n=80), titer was set to 80; and if titer was “ >6,400” (n=41), titer was set to 6,400. Antibodies titers were categorized as <10, 10 to <40, 40 to <160, 160 to <640, 640 to <2,560, and ≥2,560. Antibody titers were log-transformed for all analyses. We used a 3-parameter logistic regression model (R nplr package v0.1-7^14,15^) that, unlike traditional logistic regression, allowed some probability of infection at the highest antibody titers. Finally, we used Wilcoxon rank-sum tests to compare pre-existing antibody titers by RT-PCR or symptom status.

## RESULTS

### Cohort participation and SARS-CoV-2 pandemic waves

Between March 2020 and October 2021, we followed 2,353 people aged 0 to 94 years of age in 437 households (table S1, supplement, fig S3). Sex in the cohort is approximately equal in children but there is lower adult male participation (fig. S3, table S1). In March 2021, 90.2% of the cohort (2,123 people, table S1 and fig S1) had ELISA results.

A large initial pandemic wave occurred between March and August 2020, followed a larger second wave about a year later, consisting predominately of gamma and delta variants, from April-October 2021 (fig. S2). Between April 28, 2020 and October 14, 2021, there were 539 RT-PCR-confirmed SARS-CoV-2 infections in the cohort, with 104 (19.3%) in the first wave before August 1, 2020, 23 (4.3%) between waves, and 412 (76.4%) in the second wave after March 1, 2021.

After the first wave (October/November, 2020), 56.7% of the cohort with ELISA results was seropositive (1,130 of 1,993 people).^9^ In March 2021, 1,322 individuals were seropositive, for a seroprevalence of 62.3% (95%CI: 59.0%-65.7%, table 1 and fig S1) just prior to the second wave. Of the 1,130 people who were seropositive in October/November 2020, 1,106 (97.9%) maintained seropositivity, 20 (1.8%) were seronegative, and 4 (0.4%) did not have ELISA results in March 2021. Over the entire study period, 1,614 people (68.6% of the entire cohort of 2,353) were ever PCR-positive or seropositive.

**Table 1.** Participant characteristics

|  | Seropositive** | Seronegative | Total HICS |
| --- | --- | --- | --- |
|  | (N=1322 | (N=801 | participants with |
|  | 62.3%) | 37.7%) | ELISA |
| *All ages* |  |  | (N=2123 |
|  |  |  | 100%) |
| Age group |  |  |  |
| - 0-9y | 276 (20.9%) | 277 (34.6%) | 553 (26.0%) |
| - 10+y | 1046 (79.1%) | 524 (65.4%) | 1570 (74.0%) |
| Sex |  |  |  |
| - F | 833 (63.0%) | 464 (57.9%) | 1297 (61.1%) |
| - M | 489 (37.0%) | 337 (42.1%) | 826 (38.9%) |
| Number of PCR-positive infections* |  |  |  |
| - none | 1080 (81.7%) | 548 (68.4%) | 1628 (76.7%) |
| - one | 228 (17.2%) | 244 (30.5%) | 472 (22.2%) |
| - two | 14 (1.1%) | 9 (1.1%) | 23 (1.1%) |
|  | Seropositive | Seronegative | Total |
|  | (N=276 | (N=277 | (N=553 |
| *Ages 0-9y* | 49.9%) | 50.1%) | 100%) |
| Sex |  |  |  |
| - F | 146 (52.9%) | 138 (49.8%) | 284 (51.4%) |
| - M | 130 (47.1%) | 139 (50.2%) | 269 (48.6%) |
| Number of PCR-positive infections* |  |  |  |
| - none | 232 (84.1%) | 198 (71.5%) | 430 (77.8%) |
| - one | 42 (15.2%) | 76 (27.4%) | 118 (21.3%) |
| - two | 2 (0.7%) | 3 (1.1%) | 5 (0.9%) |
|  | Seropositive | seronegative | Total |
|  | (N=1046 | (N=524 | (N=1570 |
| *Ages 10+y* | 66.6%) | 33.4%) | 100%) |
| Sex |  |  |  |
| - F | 687 (65.7%) | 326 (62.2%) | 1013 (64.5%) |
| - M | 359 (34.3%) | 198 (37.8%) | 557 (35.5%) |
| Number of PCR-positive infections* |  |  |  |
| - none | 848 (81.1%) | 350 (66.8%) | 1198 (76.3%) |
| - one | 186 (17.8%) | 168 (32.1%) | 354 (22.5%) |
| - two | 12 (1.1%) | 6 (1.1%) | 18 (1.1%) |

NOTE: Column percentages are shown, except totals are row percentages. Characteristics of the full cohort, including those without 2021 ELISA results (which includes 21 people included in figure 4), are presented in the supplement. \*PCR-positive infections must be $\geq 60$ days apart; reported for any time during the study period;
\*\*Seropositive in March 2021

### Seropositivity and protection

Because no one in this cohort was vaccinated prior to March 2021, we could assess protection from infection-induced-immunity (and sensitivity analyses below remove the few who were vaccinated). To measure this protection, we compared the number of RT-PCR-positive infections occurring during the second wave among participants who were seropositive to those who were seronegative in March 2021 (fig. 1a). We found that infection-induced-immunity provided strong protection from infection in the second wave approximately one year later (fig. 1b and c). Protection was higher against more severe outcomes, with 64.5% protection from any infection (95% CI: 56.4%-71.1%), 69.2% protection from symptomatic infection (95% CI: 60.7%-75.9%), and 79.4% protection from moderate or severe infection (95% CI: 64.9%-87.9%).

**Fig. 1.**
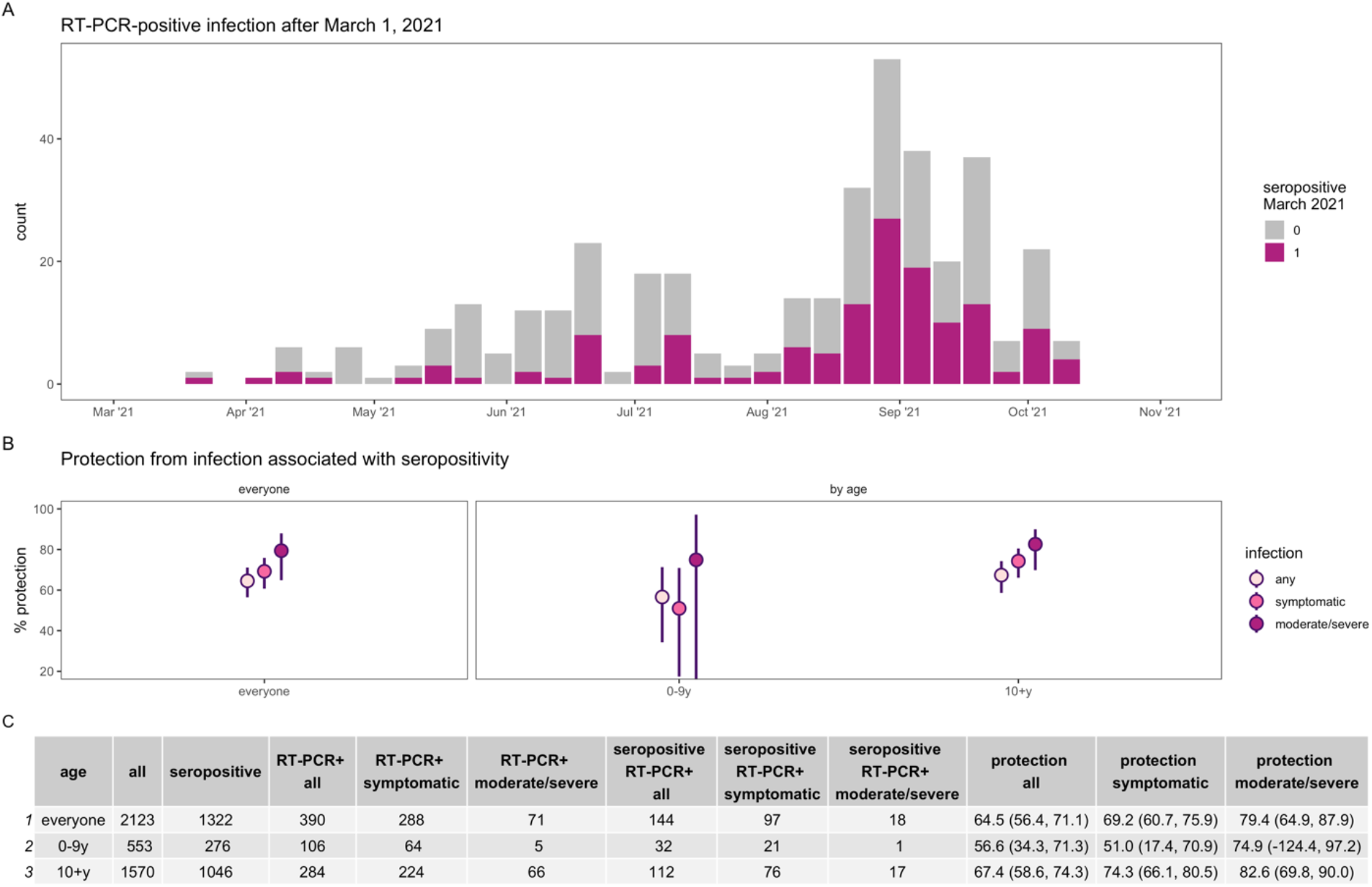
Protection from infection associated with SARS-CoV-2 seropositivity. Epidemic curve (**A**) of PCR-positive infections after March 1, 2021 colored by seropositivity status in March 2021. Percent protection (**B**) from infection (any, symptomatic, and moderate or severe) overall and by age.

Children aged 0-9 years exhibited lower protection compared to participants aged 10+ years across all severity outcomes, though there were few moderate or severe infections in the younger group. Protection against all infections was 56.6% (95% CI: 34.4%-71.3%) in ages 0-9 years and 67.4% (95% CI: 58.6%-74.3%) in ages 10+ years, and against symptomatic infection it was 51.0% (95% CI: 17.4%-70.9%) and 74.3%, (95% CI: 66.1%-80.5%), respectively.

### Antibody titers and protection

Significantly lower antibody titers were found among RT-PCR-positive (fig. 2a-c) individuals and among symptomatic (fig.2d-f) and moderate or severe (fig.2g-i) infections. However, infection and symptoms did occur in individuals with high titers, e.g., in fig. 2E.

**Fig. 2.**
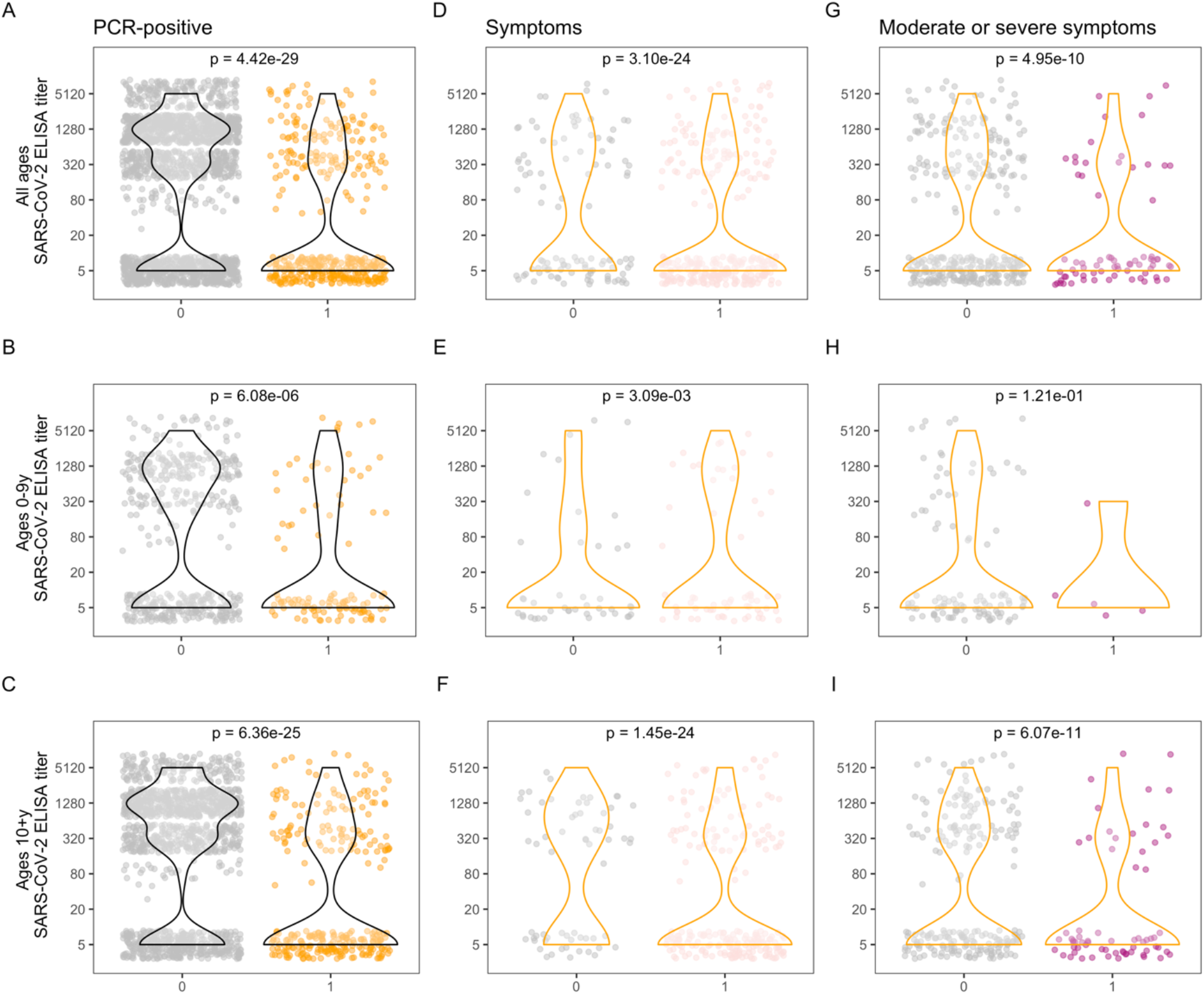
SARS-CoV-2 antibody levels and PCR and symptom positivity. Antibody levels are shown for each individual, separated by PCR-positivity **(A, B, C)**, and among PCR-positive **(D-I)**, by presence of symptoms **(D, E, F)** and presence of moderate and severe symptoms **(G, H, I)**, for all ages (top row), children 0-9 years (middle row), and ages 10+ years (bottom row). Violin plots show the density of antibody levels. P-values from Wilcoxon rank-sum tests are displayed for each comparison.

To further examine protection from SARS-CoV-2 infection, we evaluated which titers of antibodies were protective to define a correlate of protection. Antibody titers of 327 and 2,551 correlate with 50% and 80% protection from any SARS-CoV-2 infection (fig. 3a).

**Fig. 3.**
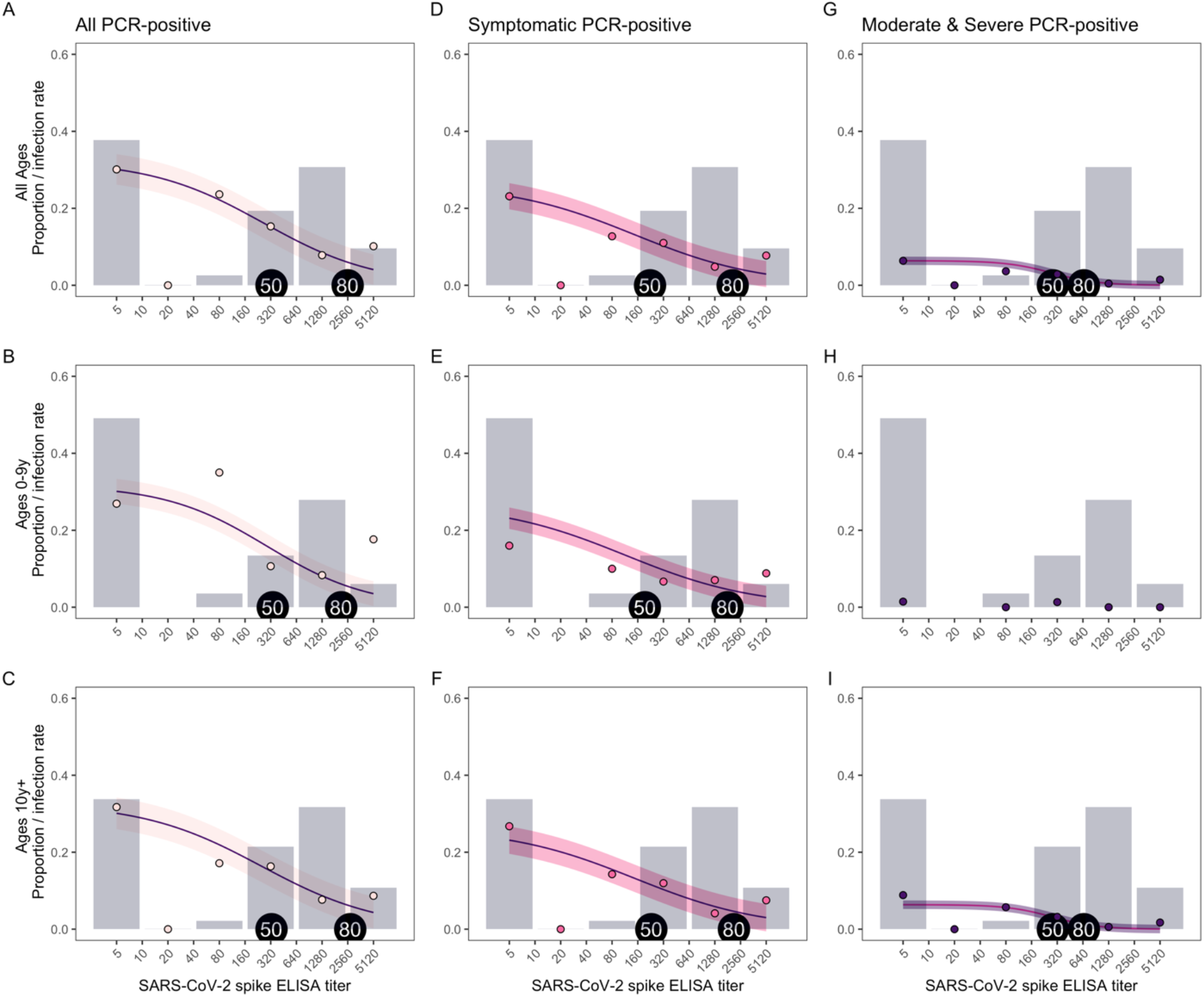
SARS-CoV-2 antibody levels needed for protection. The proportion of people with each SARS-CoV-2 spike ELISA antibody level, measured by AUC (grey bars), infection rates observed at each antibody level (colored circles), and model fits (purple lines with shaded 95% confidence intervals) are shown for all PCR-positive **(A, B, C)**, symptomatic **(D, E, F)**, and moderate and severe **(G, I)** infections, for all ages (top row), children 0-9 years (middle row), and ages 10+ years (bottom row). Because of a low number of moderate and severe infections in children ages 0-9 years (**H**), a model was not included for this group. Antibody levels associated with 50 and 80% protection are indicated with tags on each plot.

Lower antibody titers were sufficient for 50% and 80% protection against symptomatic infection (223 and 2,070, respectively, fig. 3d) and moderate or severe symptomatic infections (284 and 656, respectively, fig. 3g). Overall, a four-fold increase in antibody titers (e.g., 320 vs 80) was associated with a 29% reduction in risk of being infected (OR 0.71, 95% CI: 0.67-0.76), 30% reduction in risk of having a symptomatic infection (OR 0.70, 95% CI: 0.65-0.75), and a 35% reduction in risk of having a moderate or severe infection (OR 0.65, 95% CI: 0.56-0.75) in the second wave.

Children aged 0-9 years and participants aged 10+ years had similar antibody titers required for 50% protection, while children aged 0-9 years had somewhat lower antibody titers necessary for 80% protection (2,143 in 0-9 years vs 2,735 in ages 10+ years for all infections, fig. 3b and c; 1,700 vs 2,300 for symptomatic infections, fig. 3e and f). However, children had lower antibody titers overall (grey bars in fig. 3)—only 49.5% had titers of ≥40 and 34.0% had titers ≥640, while 66.1% of participants aged 10+ years had titers of ≥40 and 42.6% had titers ≥640. Their lower antibody levels could explain why children aged 0-9 years had lower protection associated with seropositivity compared to individuals aged 10 and older (fig. 1).

### Vaccination

Starting March 20, 2021, 61 participants (2.6%) aged 16-78 years were vaccinated. They were vaccinated with AstraZeneca/Covishield (n=47, 77.0%), Pfizer (n=1, 1.6%), and Sputnik V (n=13, 21.3%). Notably, only 6 participants were fully vaccinated with 2 vaccine doses, all of which were Sputnik V.

As sensitivity analyses, we excluded those who had received one or more vaccine doses and assessed protection associated with seropositivity and antibody levels needed for protection among those never vaccinated (figs. S4-S6). We consistently found similar protection (≤1 percentage point of difference) in estimates of seropositivity and protection (fig S4), no noticeable differences in plots of individual antibody levels (fig. S5), and similar antibody levels needed for 50% and 80% protection (fig. S6).

### Severity of first and second infections in the second wave

To assess the severity of second infections compared to first, we compared the spectrum of disease in these groups. During the second wave, there were 377 first infections and 162 second infections (following another RT-PCR-positive episode or previous seropositivity, fig. 4).

**Fig. 4.**
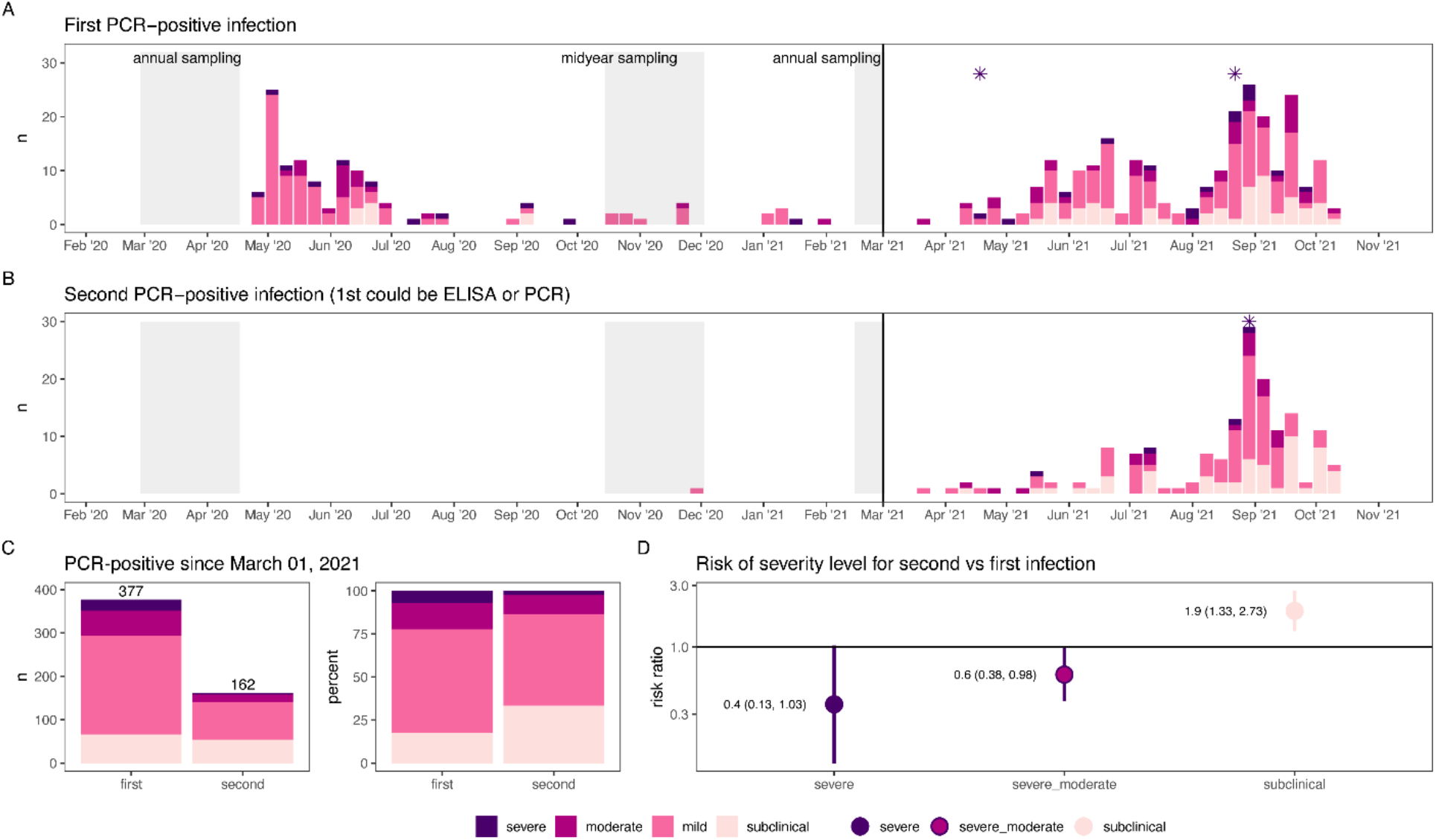
Severity of first and second SARS-CoV-2 infections in the second wave. Epidemic curves of first **(A)** and second **(B)** PCR-positive infections over the entire study period colored by severity. Second PCR-positive infections could follow a first PCR-positive infection or seropositivity determined at the midyear or 2021 annual sampling (grey shaded regions). Deaths associated with PCR-positive infections are indicated with asterisks. Severity of first and second infections was compared **(C and D)** after March 1, 2021, when variants predominated.

Second infections were less severe than first infections: moderate or severe infection was 0.6 times as likely (95% CI: 0.38-0.98) and severe infection was 0.4 times as likely (95% CI: 0.13-1.03). Second infections were also twice as likely to be subclinical (RR: 1.9, 95% CI: 1.33-2.73). There were three COVID-19 deaths following RT-PCR-positive infections, with two following first infections and one following a second infection (fig. 4).

There were no significant differences in severity between first and second infections in children aged 0-9 years (fig. S7), likely because there were fewer infections overall in young children, with few moderate or severe infections.

## DISCUSSION

In this household cohort study, we found that seropositivity was associated with protection against infection when gamma and delta SARS-CoV-2 variants predominated, determined what antibody levels were needed for protection, and showed second infections to be less severe than first infections.

Identification of antibody titers associated with protection is urgently needed^4,7^ and some measures now exist for vaccine-induced immunity,^5,6^ though not yet against all circulating variants of concern, and not stratified by severity. To date, no other studies have shown antibody titer correlates of protection from immunity induced by prior infection. From vaccine studies of Moderna mRNA-1273^5^ and Oxford AstraZenica AZD1222,^6^ looking primarily at symptomatic non-variant (mRNA-1273) or alpha (AZD1222) SARS-CoV-2 infection, 50% protection was observed even for undetectable mRNA-1273-induced neutralizing titers, and 80% protection was observed for AZD1222-induced antibodies at 264 anti-spike binding antibody units (BAU) and 506 anti-RBD BAU. While not really comparable because of antibody units, timing of antibody measurements (ours are measured much later, ∼10 months after the first wave peak), and circulating variants, the antibody titers needed for vaccine-induced antibodies to protect against symptomatic SARS-CoV-2 (original or alpha) were much lower than the prior-infection-induced antibody titers that we observed were needed to protect against primarily gamma and delta SARS-CoV-2 variants. Interestingly, AZD1222-induced antibodies were not associated with protection from asymptomatic infection,^6^ whereas we observed protection against all (including asymptomatic) infections. Additionally, studies in non-human primates suggest that vaccine-induced antibody responses are a mechanistic correlate of protection.^16-18^ A modeling study estimated that lower antibody levels are needed to protect against severe disease, which our results confirm, and that while protection will wane substantially over a year, protection from severe disease should largely remain.^19^

We previously observed 93.6% protection from symptomatic infection through March 2021 (95% CI: 51.1%-99.2%) associated with seropositivity in October/November 2020.^9^ Our current estimate of 69.2% protection from symptomatic infection is much lower. This is likely due to longer follow-up time to observe infections (and for antibody levels to wane) and the predominance of gamma and delta variants that differed from the strains against which immunity was generated.

Many studies,^9,20-28^ including several reviews,^29,30^ have demonstrated that prior infection with SARS-CoV-2 is associated with protection from reinfection, yet protection from reinfection with *variant* SARS-CoV-2 strains has not yet been established. This knowledge is needed,^25,29^ and some evidence is emerging. One study identified antibodies from four subjects that could bind and neutralize variants.^31^ A pre-print of a study in Israel (matched retrospective cohort), where vaccination is high and infection rates were <1.5%, found that prior infection was protective against reinfection while delta was predominantly circulating.^28,32^ The Israel study even found prior infection to be more protective against delta than vaccination;^28,32^ however, a recent CDC study among hospitalized patients (test-negative design) found the opposite—that vaccination was more protective than prior infection against delta.^33^ While we cannot address whether vaccination or prior infection is more protective, our findings suggest that immunity induced by prior infection is associated with moderate protection against mostly gamma and delta variants. Though lower than against the original SARS-CoV-2 strain—protection was observed in children, and across levels of severity.

Data on severity of second infections is limited, but unfortunately second infections are not universally milder^34-36^ highlighting the importance of vaccination to reduce SARS-CoV-2 burden. However, understanding the severity of repeat infections is complicated as severity differs among SARS-CoV-2 variants.^37,38^

While we did closely monitor households for SARS-CoV-2 transmission, there are likely some subclinical infections we did not detect. This means that our estimates of protection are conservative, as there would likely have been more subclinical infections in the seronegative group.

We found that seropositivity induced by prior infection is associated with protection from any SARS-CoV-2 infection (including subclinical) when mostly gamma and delta circulated, including in children. The antibody titers we presented that correlated with protection should aid in policy and control measures. Some evidence suggests that prior infection in addition to vaccination may yield strong protection.^28,39^ With the goal of reducing transmission,^40^ communities that have already suffered high infection rates will benefit greatly from vaccination, in that the added immunity from prior infection may help more than vaccination alone in reducing subclinical infections, and end the pandemic, with SARS-CoV-2 transiting to an endemic virus.

## Data Availability

Individual-level data may be shared with outside investigators following University of Michigan IRB approval. Please contact Aubree Gordon to arrange for data access.

## ACKNOWLEDGEMENTS

This work was supported by the National Institute for Allergy and Infectious Diseases at the National Institute of Health [award no. R01 AI120997 to A.G., and contract no. HHSN272201400006C to A.G.], and a grant from Open Philanthropy.

We are extremely grateful to the families who participated in this study and to the incredibly dedicated teams at the Centro de Salud Sócrates Flores Vivas, the Nicaraguan National Virology Laboratory at the Nicaraguan Ministry of Health and the Sustainable Sciences Institute who worked through this pandemic. We would like to thank Leo Poon for providing the protocol and controls for RT-PCR testing, and Florian Krammer for sharing RBD and Spike constructs as well as technical advice. We are grateful to Janet Smith, Melanie Ohi and their groups at the Center of Structural Biology at the UM Life Sciences Institute for producing proteins and antibodies for the ELISAs.

**PSP Study Group members (all from Mount Sinai):

Ana Silvia Gonzalez-Reiche

Zain Khalil

Adriana van de Guchte

Jian Zhang

Keith Farrugia

Hala Alshammary

Aria Rooker

Christian Cognigni

Viviana Simon

