## Supplement for "An immune correlate of SARS-CoV-2 infection and severity of reinfections"

^5^Mount Sinai

^6^Division of Infectious Diseases and Vaccinology, School of Public Health, University of California, Berkeley, California, USA

* corresponding author

**Corresponding author:**

Aubree Gordon, Department of Epidemiology, School of Public Health, University of Michigan, Ann Arbor, 5622 SPH I, 1415 Washington Heights, Ann Arbor, MI 48109-2029; Tel. 1-510-409- 5495;

**Table of Contents**

**Material and Methods**

TCOVID Transmission sub-study

Symptoms

Severity classification

Age standardization

Sequencing

**Results**

Cohort participation

Age standardization

**Figures**

Fig. S1.

Fig. S2.

Fig. S3.

Fig. S4.

Fig. S5.

Fig. S6.

Fig. S7.

**Tables**

Table S1.

**Materials and Methods**

TCOVID transmission sub-study

When a participant in HICS met the testing criteria and tested positive at the study health clinic for SARS-CoV-2 by an initial rapid test or real-time reverse transcription polymerase chain reaction (RT-PCR), their household was invited to participate in the TCOVID sub-study and an intensive monitoring period began. Activated TCOVID households were visited by study staff on approximately days 0, 3, 7, 14, 21, and 30. Respiratory samples and daily symptom diaries were collected during home visits.

Household activations did not start immediately when the first SARS-CoV-2 cases were identified by RT-PCR due to administrative delays. The first RT-PCR-positive infection in this study occurred on April 28, 2020, while the first household activation started on June 18^th^ (fig. S1). There were 159 households activated, with 27 activated twice, and one household activated three times.

### Symptoms

Participants whose households participated in the transmission sub-study had daily symptom diaries from intensive monitoring periods.

Additionally, data on COVID-19 signs and symptoms were systematically collected at each medical visit, some of which were home visits. At each medical visit, all patients were asked about standard symptoms, and suspected COVID-19 cases were asked about an additional set of COVID-specific questions. Symptoms reported in medical visits within 14 days prior to and 27 days after the first RT-PCR-positive test or symptom onset were included for evaluation of severity.

There were 1,846 total clinic records occurring within 14 days prior to and 27 days after the first RT-PCR positive test or onset of symptoms. These records consisted of, per infection, up to 5 visits to the clinic, 15 home visits for transmission sub-study participants, and 8 home visits for participants not currently participating in intensive monitoring.

Between clinic records and intensive monitoring period diaries, symptom data was available for all RT-PCR-positive infections.

### Severity classification

Infection severity was classified similarly to previously (Maier, Kuan, Saborio et al) though using symptom diaries and clinic record exclusively and not the COVID questionnaire administered in October 2020. There were three COVID-19-related deaths among PCR-positive infections and five total, identified and classified through death certificates and clinic records. COVID-19-related deaths and hospitalizations were classified as severe. Lower respiratory symptoms were classified as moderate; these included difficulty or rapid breathing, chest pain or tightness in chest, crepitus, chest wall indrawing, rhonchi, wheezing, and overall poor condition. Loss of smell/taste or at least two symptoms were classified as mild. The presence of only one symptom was classified as subclinical.

Age standardization

Severity proportions among PCR-positive infections were age-standardized to 1) the entire HICS study population, 2) Nicaragua, and 3) the USA, using data from the United Nations.

Sequencing

To identify the SARS-CoV-2 variants circulating over time, sequencing was performed on 481 samples (157 at the Nicaraguan National Virology Lab and 324 at Mount Sinai in New York).

*Nicraraguan National Virology Lab*

SARS-CoV-2-positive samples from 157 cases, tested at Virology laboratory in Centro Nacional de Diagnóstico y Referencia in Managua, Nicaragua, were retrieved from storage and included in this study. All specimens were nasopharyngeal swabs originating from patients of the local health care centers of Nicaragua.

Specimens underwent RNA extraction using the QIAGEN QIAamp^(R)^ Viral RNA Mini kit. RNA was reverse-transcribed then amplified using multiplexed PCR of 98 × ~400 bp amplicons that enabled evaluation of ~95% of the SARS-CoV-2 genome. Amplicons were prepared for nanopore using the ONT Native Barcoding Expansion kit (EXP-NBD196). The kit was used according to the manufacturer’s protocol. Up to 48 samples were multiplexed on FLO-MIN106 flow cells (R9.4.1 flow cells) version and sequenced on a MinION Mk1B device. The *RAMPART* (v1.0.6) software package^1^ was used to monitor sequencing performance in real-time across all amplicons. The run was terminated and the flow-cell washed using the ONT Flow Cell Wash kit (EXP-WSH004), allowing re-use in subsequent runs.

The resulting reads were basecalled using *Guppy* as integrated in to MinKnow (21.06.13) and aligned to the Wuhan-Hu-1 reference genome ([MN908947.3](https://www.ncbi.nlm.nih.gov/nuccore/MN908947.3)). Porechop tool for finding and removing adapters from [Oxford Nanopore](https://nanoporetech.com/) reads was used to trim primer sequences from the termini of read alignments. Consensus-level variant candidates were identified using the workflows developed by ARCTIC using Medaka (0.11.5) to variants detection.

*Mount Sinai*

Sequencing was performed as previously described.^2^

1.	Mapleson D, Drou N, Swarbreck D. RAMPART: a workflow management system for de novo genome assembly. Bioinformatics 2015;31(11):1824-6. DOI: 10.1093/bioinformatics/btv056.

2.	Gonzalez-Reiche AS, Hernandez MM, Sullivan MJ, et al. Introductions and early spread of SARS-CoV-2 in the New York City area. Science 2020;369(6501):297-301. DOI: 10.1126/science.abc1917.

**Results**

Cohort participation

Between March 2020 and October 2021, 72 people enrolled and 139 people left the cohort. Participants consisted of 596 (25.3%) children under 10 years of age and 1,757 (74.7%) people 10 years of age and older (a population pyramid is shown in fig. S3).

Age standardization

Severity proportions (fig. S7) were similar when standardized to the age distributions of the full HICS cohort, Nicaragua, and the USA. Though, expectedly, severity was higher for the USA with an older population.

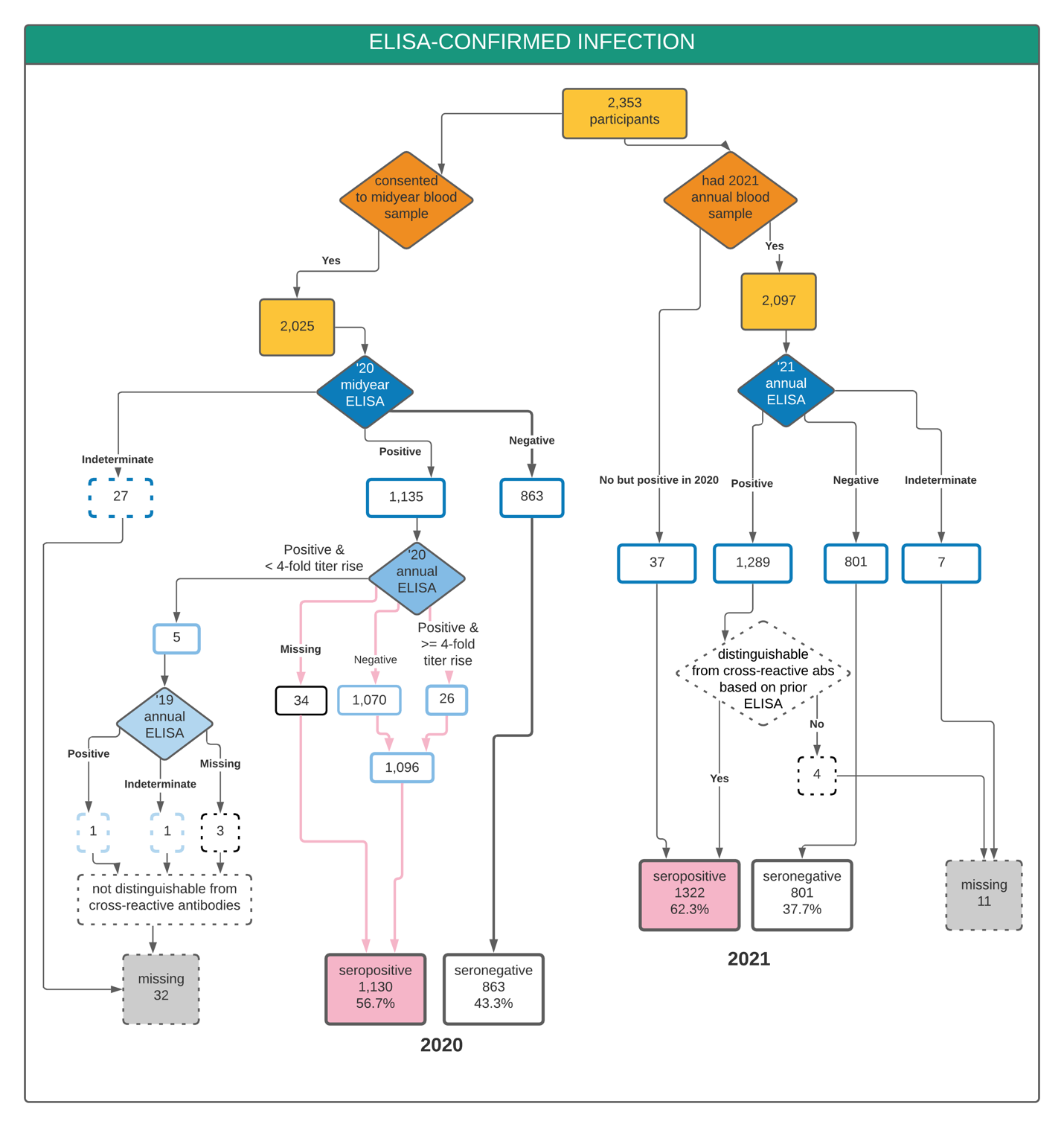

**Fig. S1. Flowchart of ELISA results and classification of serostatus.**

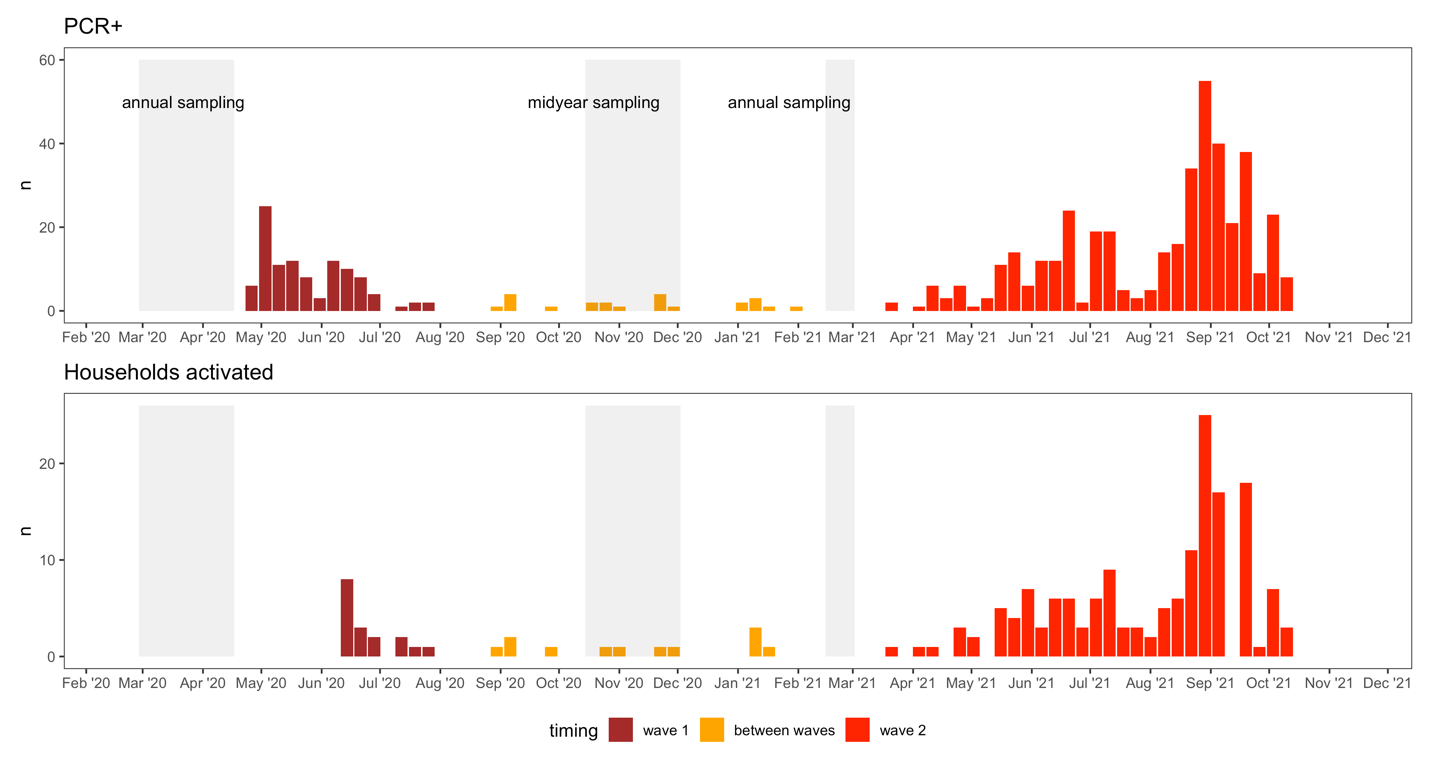

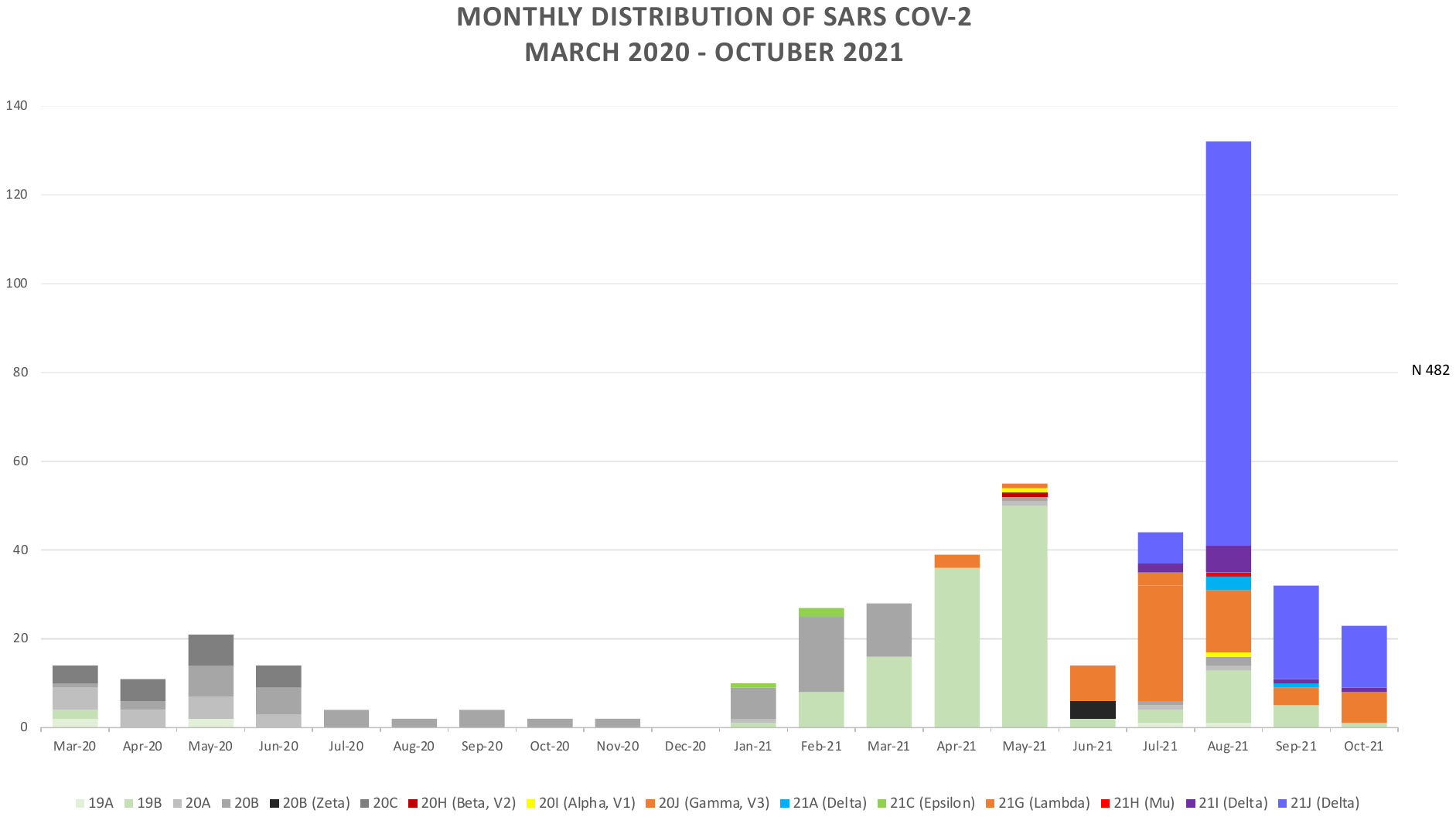

**Fig. S2. Epidemic and household activation timing and sequencing.** Sequences are from 481 samples from Nicaragua, including but not exclusively from the cohort.

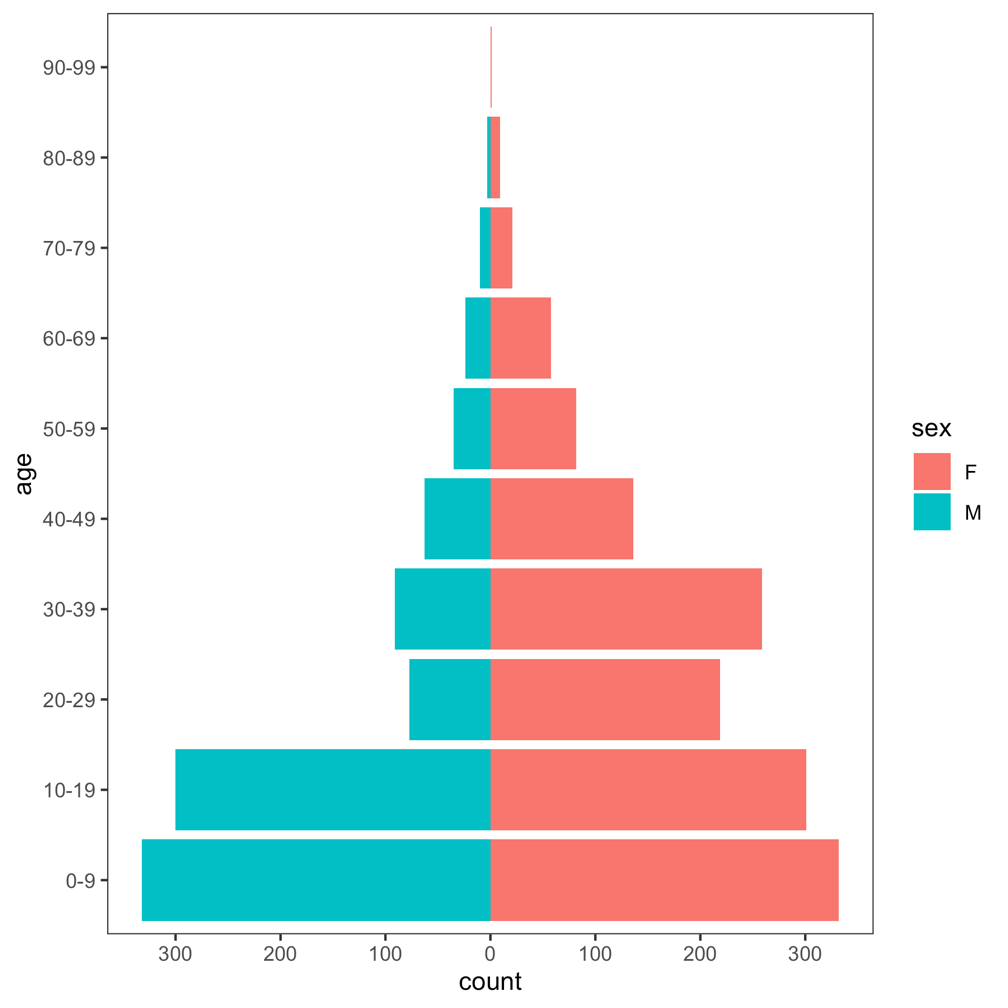

**Fig. S3. Age pyramid for HICS population.**

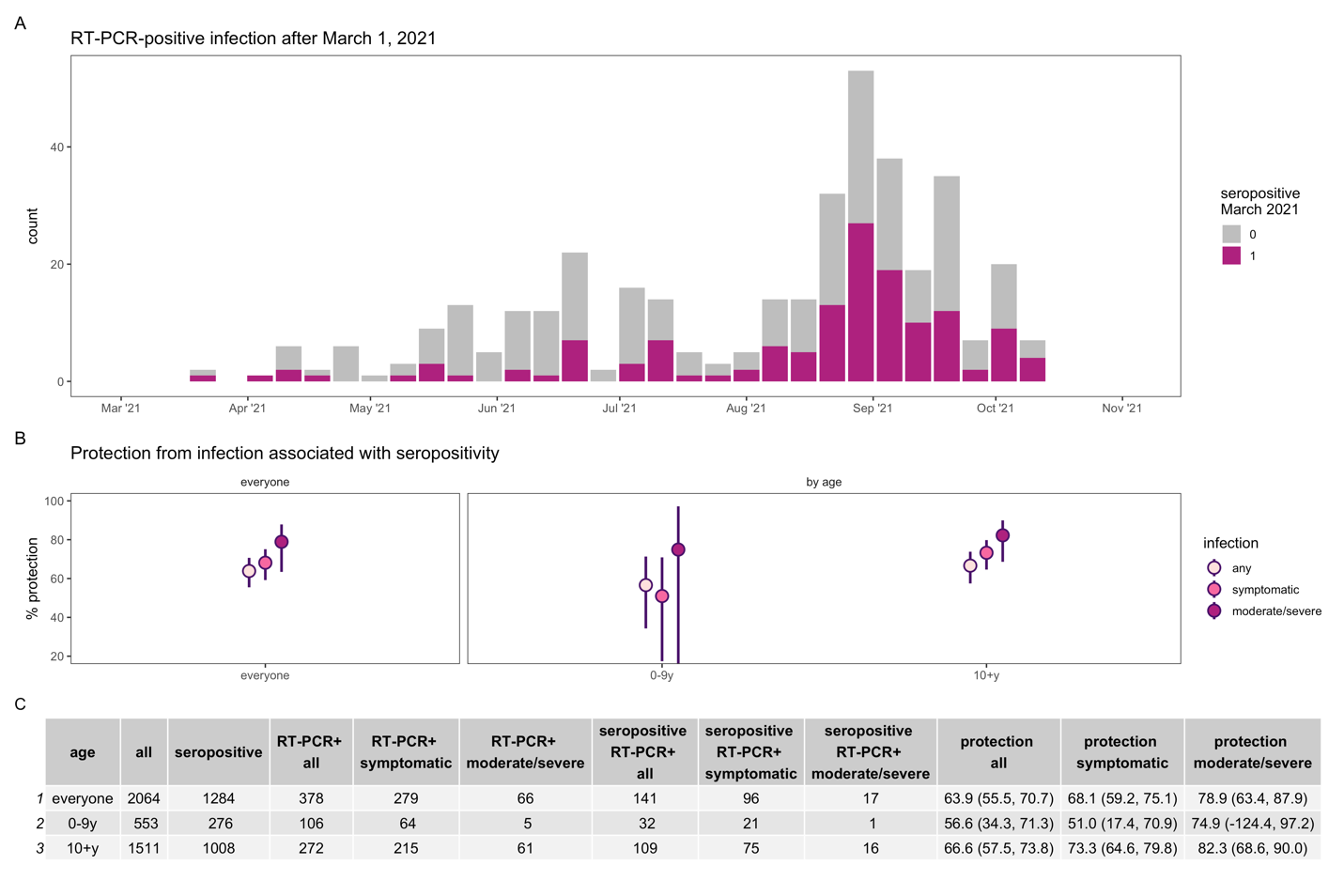

**Fig. S4. Protection from infection associated with SARS-CoV-2 seropositivity, excluding 59 participants 16 years of age and older who were vaccinated starting in March 2021.** Epidemic curve (**A**) of PCR-positive infections after March 1, 2021 colored by seropositivity status in March 2021. Percent protection (**B**) from infection (any, symptomatic, and moderate or severe) overall and by age.

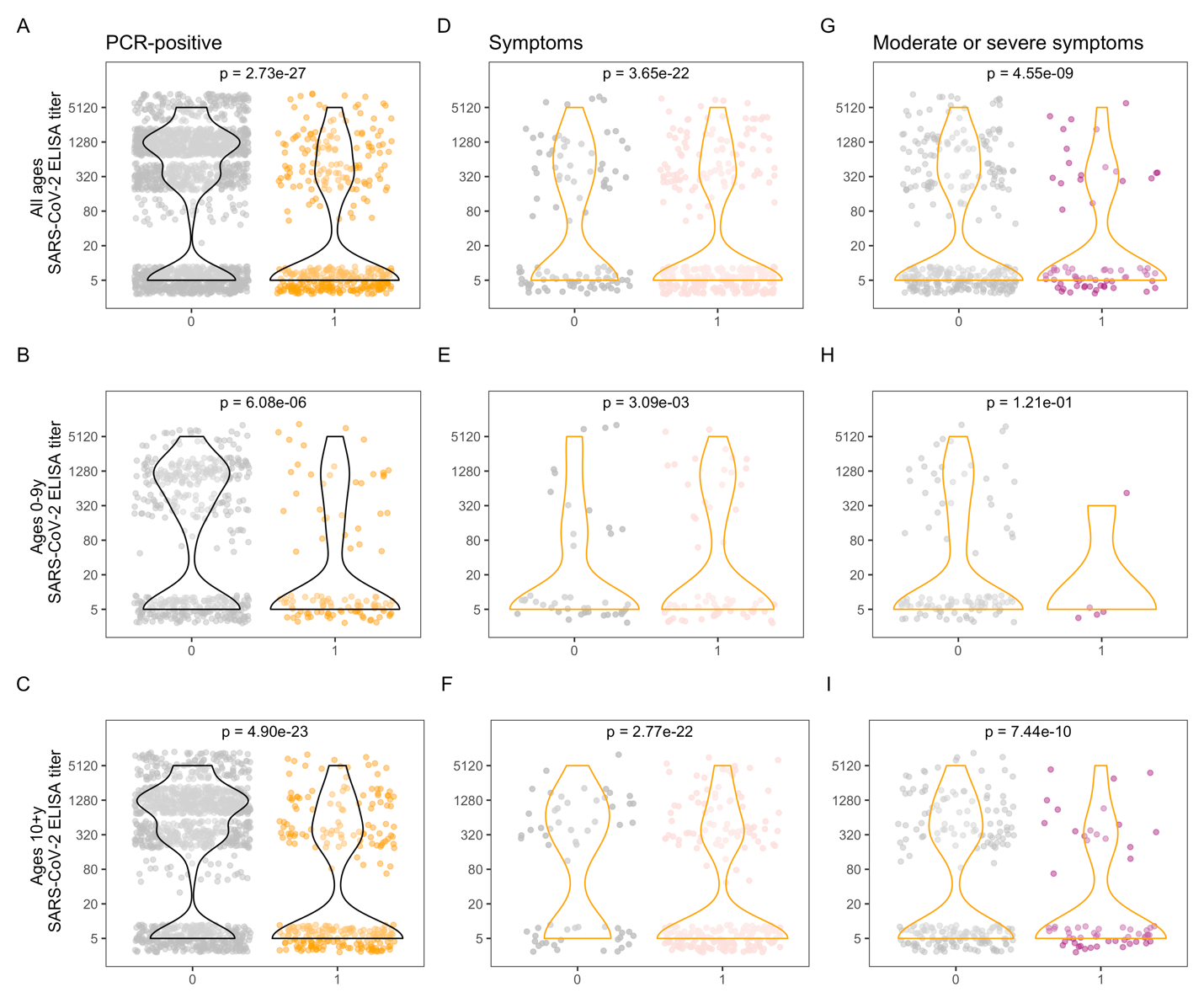

**Fig. S5. SARS-CoV-2 antibody levels and PCR and symptom positivity, excluding 59 participants 16 years of age and older who were vaccinated starting in March 2021.** Antibody levels are shown for each individual, separated by PCR-positivity **(A, B, C)**, and among PCR-positive **(D-I)**, by presence of symptoms **(D, E, F)** and presence of moderate and severe symptoms **(G, H, I)**, for all ages (top row), children 0-9 years (middle row), and ages 10+ years (bottom row). Violin plots show the density of antibody levels. P-values from Wilcoxon rank-sum tests are displayed for each comparison.

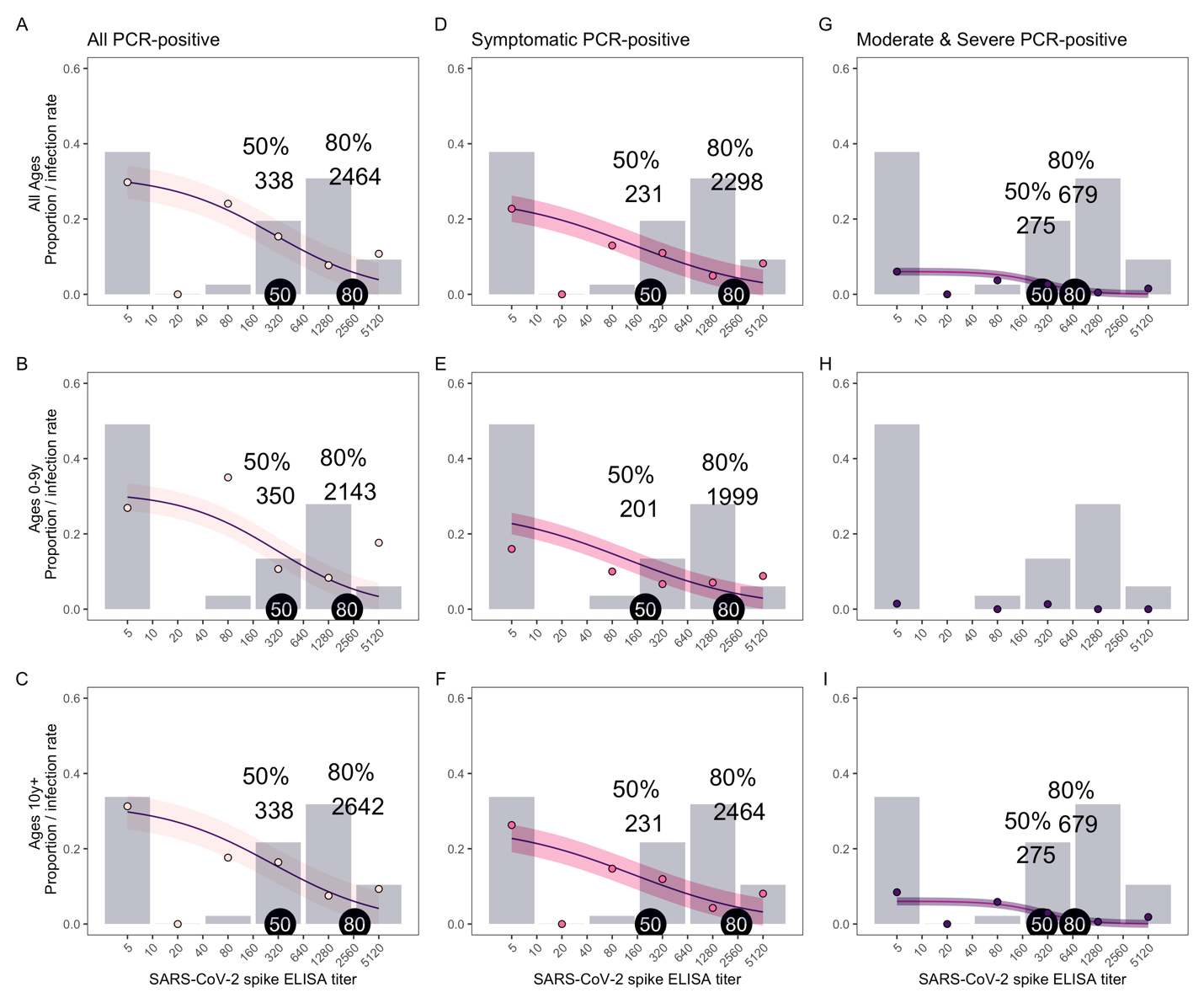

**Fig. S6. SARS-CoV-2 antibody levels needed for protection, excluding 59 participants 16 years of age and older who were vaccinated starting in March 2021.** The proportion of people with each SARS-CoV-2 spike ELISA antibody level, measured by AUC (grey bars), infection rates observed at each antibody level (colored circles), and model fits (purple lines with shaded 95% confidence intervals) are shown for all PCR-positive **(A, B, C)**, symptomatic **(D, E, F)**, and moderate and severe **(G, H, I)** infections, for all ages (top row), children 0-9 years (middle row), and ages 10+ years (bottom row). Because of a low number of moderate and severe infections in children ages 0-9 years, a model was not included for this group. Antibody levels associated with 50 and 80% protection are indicated with tags on each plot.

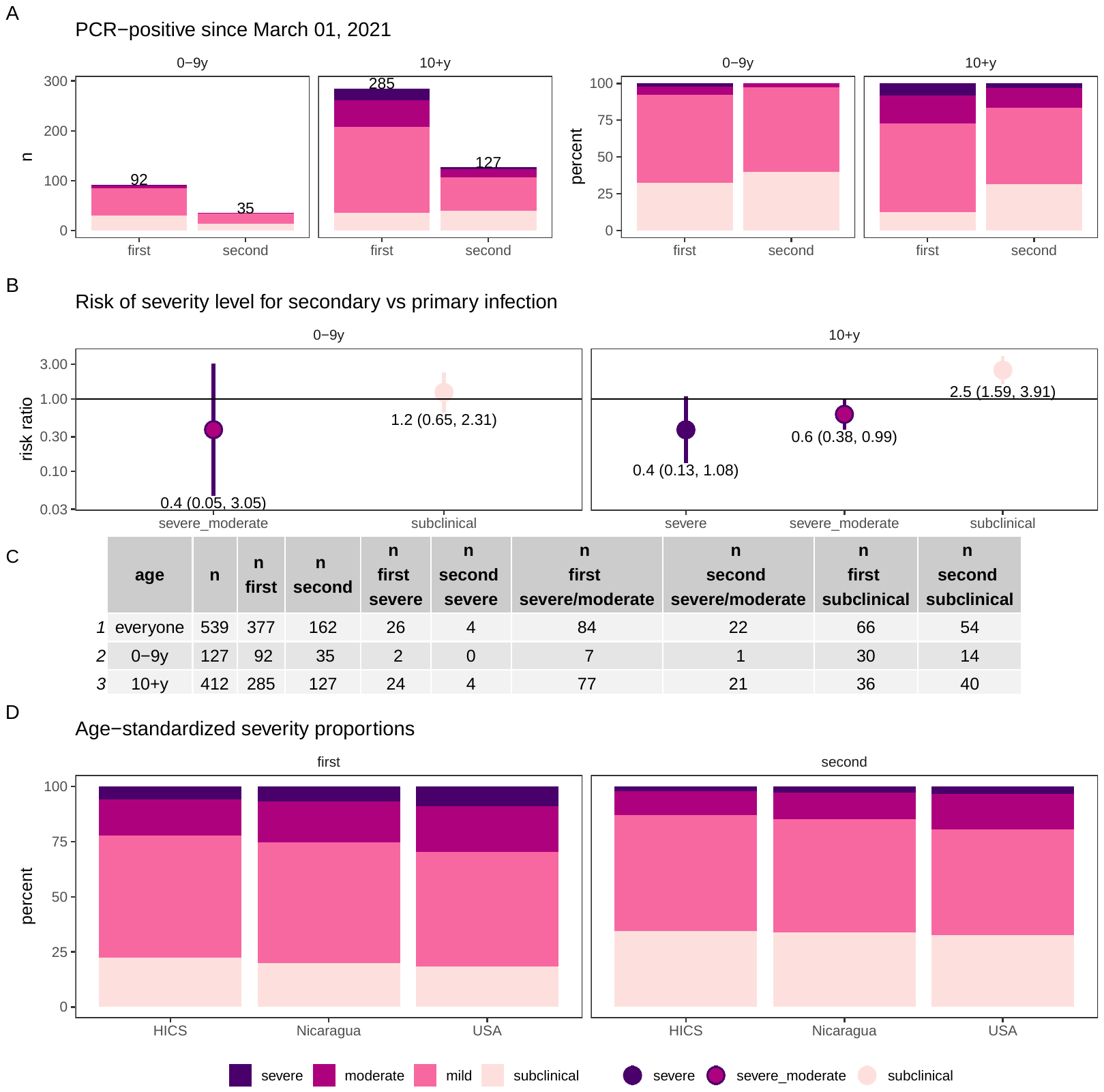

**Fig. S7. Severity of first and second SARS-CoV-2 infections, by age group.** Second PCR-positive infections could follow a first PCR-positive infection or seropositivity determined at the midyear or 2021 annual sampling. Severity of first and second infections after March 1, 2021, when variants predominated, by age (**A** and **B**). Table of infections by severity and age (**C).** Age-standardized severity proportions (**D**), standardized to the full HICS cohort, Nicaragua, and the USA.

| Table S1. Participant characteristics of all HICS | | | |
| --- | --- | --- | --- |
| *All ages* | has 2021 ELISA (N=2123) | no 2021 ELISA data  (N=230) | Total HICS participants  (N=2353) |
| Age group |  |  |  |
| - 0-9y | 553 (26.0%) | 43 (18.7%) | 596 (25.3%) |
| - 10+y | 1570 (74.0%) | 187 (81.3%) | 1757 (74.7%) |
| Sex |  |  |  |
| - F | 1297 (61.1%) | 121 (52.6%) | 1418 (60.3%) |
| - M | 826 (38.9%) | 109 (47.4%) | 935 (39.7%) |
| Number of PCR-positive infections* |  |  |  |
| - none | 1628 (76.7%) | 209 (90.9%) | 1837 (78.1%) |
| - one | 472 (22.2%) | 21 (9.1%) | 493 (21.0%) |
| - two | 23 (1.1%) | 0 (0.0%) | 23 (1.0%) |
| *Ages 0-9y* | has 2021 ELISA (N=553) | no 2021 ELISA data  (N=43) | Total (N=596) |
| Sex |  |  |  |
| - F | 284 (51.4%) | 18 (41.9%) | 302 (50.7%) |
| - M | 269 (48.6%) | 25 (58.1%) | 294 (49.3%) |
| Number of PCR-positive infections* |  |  |  |
| - none | 430 (77.8%) | 39 (90.7%) | 469 (78.7%) |
| - one | 118 (21.3%) | 4 (9.3%) | 122 (20.5%) |
| - two | 5 (0.9%) | 0 (0.0%) | 5 (0.8%) |
| *Ages 10+y* | has 2021 ELISA (N=1570) | no 2021 ELISA data  (N=187) | Total (N=1757) |
| Sex |  |  |  |
| - F | 1013 (64.5%) | 103 (55.1%) | 1116 (63.5%) |
| - M | 557 (35.5%) | 84 (44.9%) | 641 (36.5%) |
| Number of PCR-positive infections* |  |  |  |
| - none | 1198 (76.3%) | 170 (90.9%) | 1368 (77.9%) |
| - one | 354 (22.5%) | 17 (9.1%) | 371 (21.1%) |
| - two | 18 (1.1%) | 0 (0.0%) | 18 (1.0%) |
| *PCR-positive infections must be >=60 days apart; reported for any time during the study period | | | |
